# SUNSCREEN EFFICACY AGAINST UVA1- AND VISIBLE LIGHT-INDUCED SKIN PIGMENTATION IS INFLUENCED BY ETHNICITY

**DOI:** 10.64898/2025.12.16.25342374

**Authors:** Florian Dimmers, Niklas Lück, Yimeng Wang, Christian Staerk, Tao Zhang, Jean Krutmann

## Abstract

**Background:** There is growing evidence that individuals with different skin phototypes require tailored approaches to achieve optimal photoprotection. Individuals with darker skin phototypes are more prone to UVA1- and visible light-induced pigmentation, whereas lighter phototypes are more susceptible to shorter wavelengths such as UVB and UVA2. Thus, skin phototype is an important determinant of sunscreen efficacy. In the present study we have asked if ethnicity – independent of phototype - is another factor affecting sunscreen efficacy.

**Objectives:** (i) To determine the overall photoprotective effects of two sunscreen formulations against UVA1, visible light (VL), and combined visible light *plus* UVA1, and (ii) to compare the photoprotective efficacy of both products between Han Chinese and Caucasian participants.

**Methods:** Forty healthy volunteers (N=20 Han Chinese; N=20 Caucasian), matched for phototype, constitutive pigmentation, gender, and age, were exposed to VL, UVA1, and combined VL *plus* UVA1 to induce pigmentation responses following standardized irradiation protocols. Skin responses across treated and untreated sites were analysed using linear mixed-effects models.

**Results:** Across all participants, both sunscreen formulations provided significant protection against VL, UVA1, and combined VL *plus* UVA1. Notably, photoprotective efficacy against UVA1-induced immediate (IPD) and persistent pigment darkening (PPD) differed significantly between ethnic groups, with one formulation showing stronger protection in Han Chinese.

**Conclusion:** This study indicates that ethnicity could influence sunscreen efficacy. Thus, sunscreens should not only be tailored to different phototypes, but also consider ethnic background.

## INTRODUCTION

Individual susceptibility to solar radiation depends on both wavelength and skin phototype. Fair-skinned individuals (Fitzpatrick^1^ phototype I-II) are particularly vulnerable to UVB-induced erythema and photocarcinogenesis^2^, whereas darker phototypes (III-IV) show stronger pigmentation responses to long-wave UVA1 and visible light (VL)^3–6^, likely due to intrinsic differences in pigment biology^7,8^.

As a result, photoprotection strategies increasingly address the specific vulnerability profiles of different phototypes.

Recent clinical studies have demonstrated that UVA1 (340-400 nm) and high-energy visible (HEV) light (400-440 nm) can induce persistent pigmentation in darker skin phototypes and contribute to skin pigmentary disorders such as melasma and post-inflammatory hyperpigmentation^9–11^. Protection against VL is limited because standard sunscreens are primarily designed to block UVB and UVA2. Historically, sunscreen performance has been evaluated through UVB-driven SPF testing in fair-skinned individuals, which does not capture efficacy against UVA1 or VL. As a result, many commercially available sunscreens offer inadequate protection in these wavelength ranges^12^.

Although skin phototype strongly influences cutaneous responses to solar radiation, ethnicity – independent of phototype – may also contribute. Han Chinese individuals, predominantly within Fitzpatrick III-IV^13,14^, are more prone to pigmentary disorders as a major manifestation of photoaging^15,16^, whereas wrinkle formation predominates in Caucasians^17^.

To date, no controlled study has systematically compared UVA1- and VL-induced pigmentation responses between Han Chinese and Caucasian individuals while evaluating the efficacy of sunscreen formulations in both groups.

This study addresses this knowledge gap by (i) determining the overall photoprotective effects of two sunscreen formulations against UVA1, VL and combined VL *plus* UVA1, and (ii) comparing their efficacy between Han Chinese and Caucasian participants.

## MATERIALS AND METHODS

### Study design

This monocentre, interventional, exploratory *in vivo* study was designed to assess the photoprotective efficacy of two sunscreen formulations, with a particular focus on potential differences between ethnic groups.

All study procedures were conducted under standardised conditions and in accordance with the Declaration of Helsinki and the principles of Good Clinical Practice (ICH-GCP) at the study centre of the IUF-Leibniz Research Institute for Environmental Medicine, Düsseldorf (Germany), between 23 May and 12 December 2024. The study protocol was approved by the Ethics Committee of the Medical Faculty of the Heinrich Heine University Düsseldorf (Approval number: 2023-2713; Date: 14 May 2024) and was registered in the German Clinical Trials Register (DRKS; ID: DRKS00034235; registered on 16 May 2024). Written informed consent was obtained from each participant prior to study inclusion.

### Study subjects

A total of 40 healthy volunteers aged 20-42 years (mean age: 26.65 ± 5.1 years) were enrolled (n=20 Caucasian; n=20 Han Chinese). Both genders were included. Female participants of childbearing potential had to provide a negative urine pregnancy test at screening. The two ethnic groups were matched for gender, age, and constitutive pigmentation (Individual Typology Angle (ITA°)^18^ at baseline) to ensure comparability between ethnic groups. One Han Chinese participant discontinued after visit 3 due to subjective discomfort during irradiation. No unexpected skin reactions to visible light exposure occurred in any participant.

Eligibility criteria included good general health, absence of photosensitivity, and no relevant skin disease. Further requirements were a normal diet (excluding vegetarian or vegan diets), use of contraception in female participants, no sunbed use or extensive sun exposure within three months before enrolment and during the trial, and no intake of nutritional supplements.

Key exclusion criteria comprised pregnancy and breastfeeding, smoking, use of oral antioxidants or vitamins within three months before enrolment, known allergies or intolerances to the components of the investigational products.

### Study procedures

The study comprised of eight visits (Figure 1A). After a seven-day washout period, at visit 1, the skin phototype according to Fitzpatrick was determined and the baseline pigmentation was quantified to allow matching between ethnic groups.

**Figure 1.**
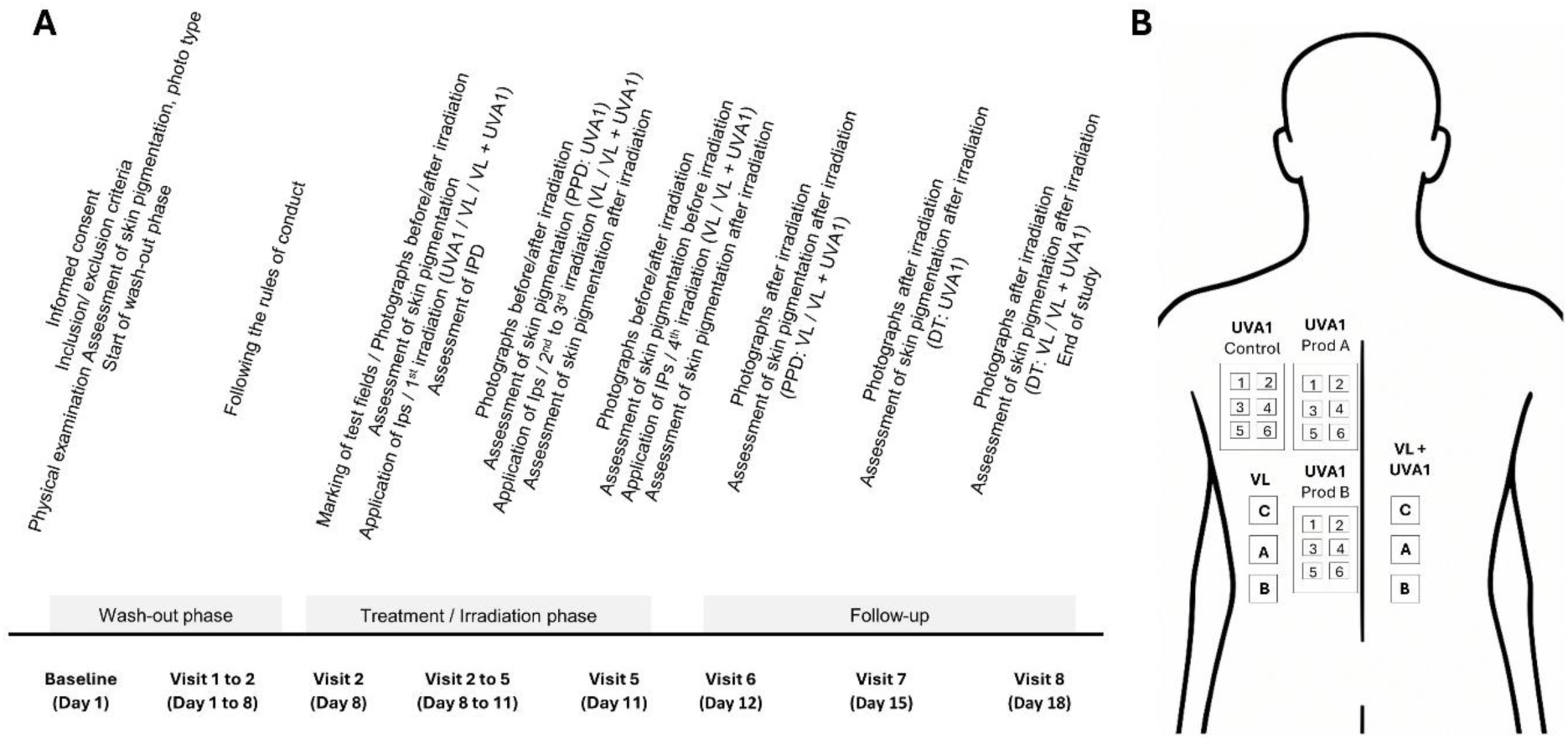
Study procedures for each subject (A) and schematic illustration of irradiation fields of one participant (B). (A) Overview of the study procedure that each participant underwent. VL, visible light; IP, Investigational Product; IPD, Immediate Pigment Darkening; PPD, Persistent Pigment Darkening; DT, Delayed Tanning. (B) Schematic illustration of the irradiation fields. C indicates the control field, A denotes the test fields treated with product A, and B denotes the test fields treated with product B. Numbers 1-6 indicate the different UVA1 doses, with 1 corresponding to the highest dose (100 J/cm²) and 6 to the lowest dose (32 J/cm²).

**Figure 2.**
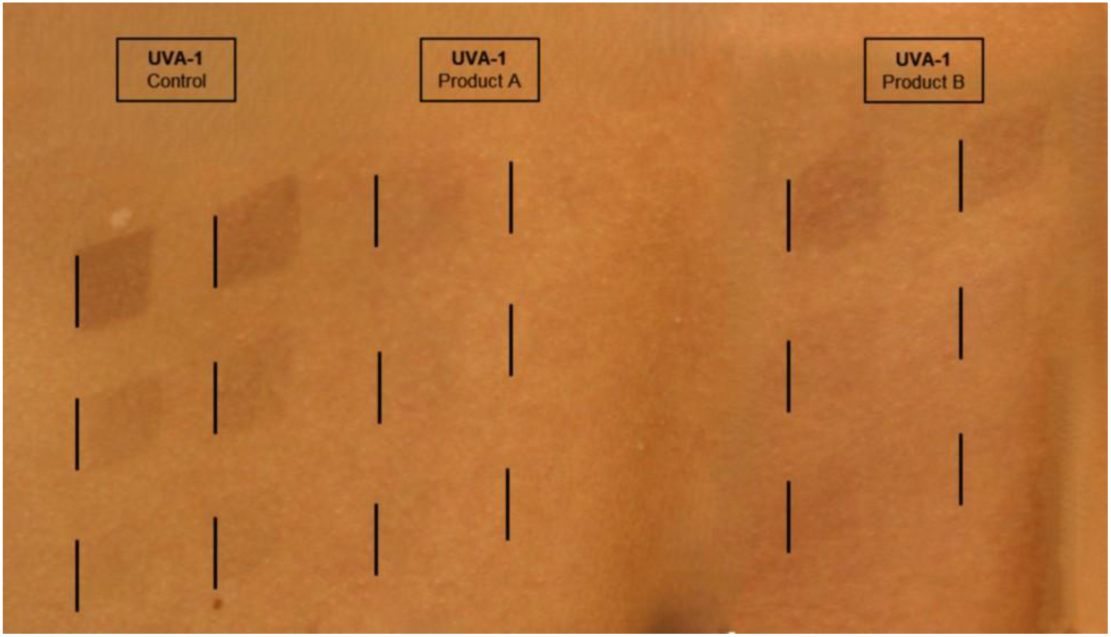
Exemplary clinical image of immediate pigment darkening (IPD) after UVA1 exposure in control and product-treated fields (Product A and B). Representative clinical photograph illustrating immediate pigment darkening (IPD) after UVA1 exposure. The image displays the control field and the two treatment fields (product A and B). The six black markers indicate the six different UVA1 doses applied to each field. For improved visual comparability, the irradiation fields were digitally rearranged.

**Figure 3.**
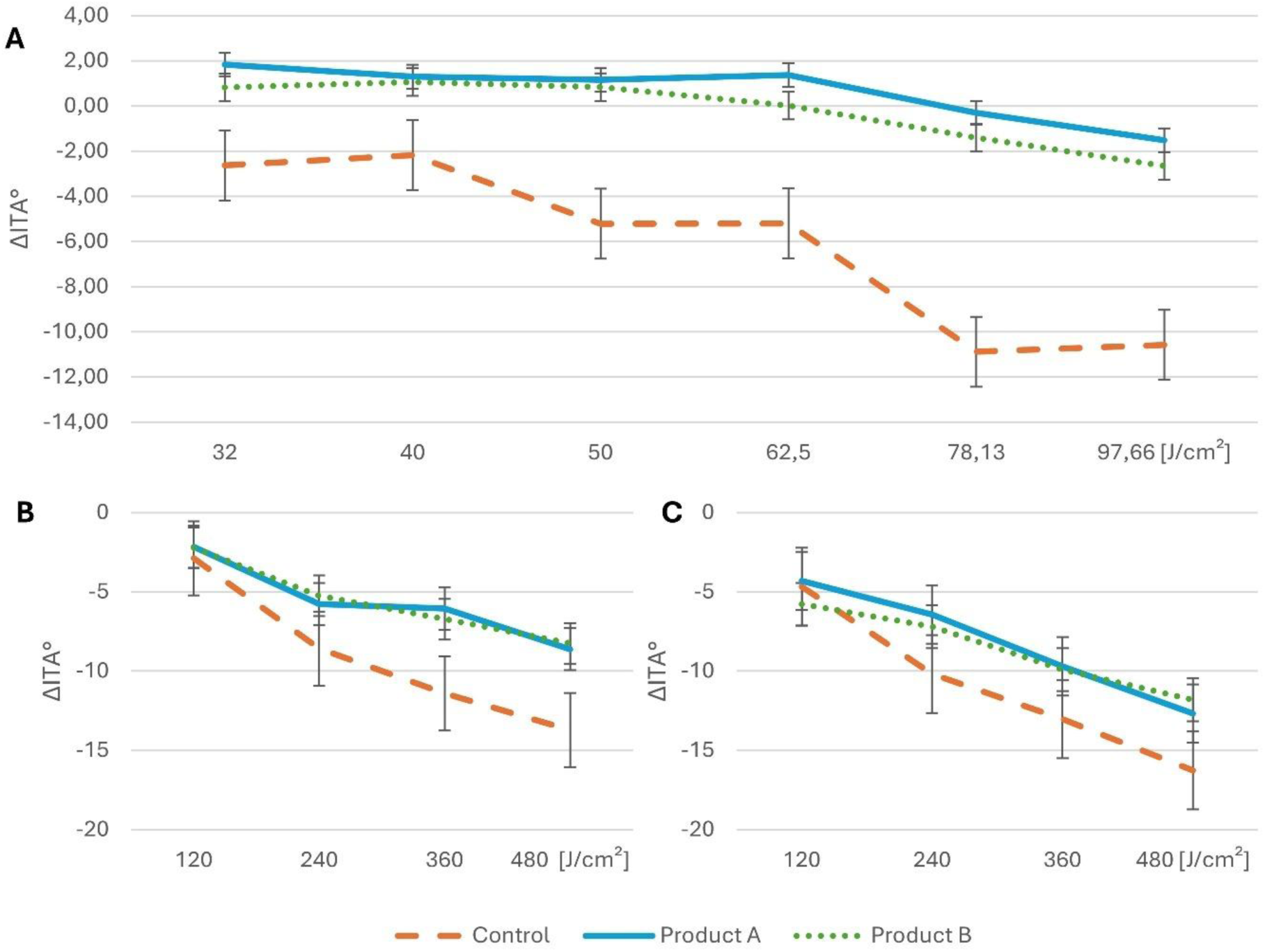
Dose-dependent changes in immediate pigment darkening (IPD; ΔITA°) in control and treatment fields after exposure to (A) UVA1, (B) visible light, (C) VL *plus* UVA1. Graphs showing mean changes in Individual Typology Angle (ΔITA°) in control and treatment fields (product A and product B). Data are displayed separately for (A) UVA1, (B) VL, and (C) VL plus UVA1. Data are presented as means ± standard error of means (SEM).

During visit 2 to visit 5, the interventions were carried out. The UVA1 irradiation was performed once at visit 2 using a Sellamed 2000 UVA1 partial therapy system (UVA1: 340 – 400 nm) (Sellas Medizinische Geräte GmbH, Ennepetal, Germany). The participants were exposed to six predefined UVA1 doses (approx. 100 J/cm², 78 J/cm², 62 J/cm², 50 J/cm², 40 J/cm², and 32 J/cm²). These doses were applied to one control and two treatment sites. Each irradiation site consisted of six irradiation fields of approx. 1 x 1 cm, with one UVA1 dose applied per field (see Figure 1B). The VL and the combined visible light *plus* UVA1 irradiations were performed on four consecutive days (visit 2 to visit 5) with an irradiance of approx. 100 mW/cm² using a Sellas vis400 irradiation device (visible light: 400 – 440 nm) (Sellas Medizinische Geräte GmbH, Ennepetal, Germany). For irradiation with visible light *plus* UVA1 (visible light *plus* UVA1: 390 – 440 nm), the standard built-in GG400 filter (Schott AG, Mainz, Germany) was removed. VL and VL *plus* UVA1 were each applied to a separate set of one control and two treatment fields (each approx. 3 x 3 cm). The same fields were irradiated across all four visits with a dose of 120 J/cm² per day, resulting in a cumulative dose of 480 J/cm². The described irradiation scheme is consistent with currently proposed practices^19^. The corresponding irradiation spectra of the different light sources can be found in Figure S1 of the Supporting Information.

The skin pigmentation was determined by a Minolta CR-400 chroma meter (Konica Minolta Inc., Osaka, Japan), and a DSM III colorimeter (Cortex Technology, Hadsund, Denmark).

### Product application and safety

At visit 2 to 5, the two sunscreen formulations (products A and B) were applied to the designated treatment fields at a dosage of 2 mg/cm² before irradiation. Compliance with restrictions and adverse events were systematically assessed at all visits. No product-related adverse events occurred.

### Statistical Analysis

The main objective of the study was to evaluate potential differences in the protective efficacy of two sunscreen formulations between Han Chinese and Caucasian participants following UVA1, VL, and VL *plus* UVA1 exposure, as determined by variations in immediate pigment darkening (IPD), persistent pigment darkening (PPD), or delayed tanning (DT) values. Furthermore, different sunscreen-free tanning behaviour between the ethnicities and overall product efficacies were investigated. Due to the multitude of experimental conditions and research questions, the following analyses are exploratory in nature. Therefore, p-values are reported descriptively and were not adjusted for multiple testing; findings should be interpreted in conjunction with effect sizes and 95% confidence intervals^20^.

Statistical analyses were performed using R version 4.4.1^21^. For each combination of endpoint (change in ITA° and MIndex), irradiation type (UVA1, VL, and VL *plus* UVA1), and tanning kinetic (immediate pigment darkening (IPD), persistent pigment darkening (PPD), and delayed tanning (DT)) a separate linear mixed-effect model^22^ (R packages Ime4^23^ and emmeans^24^) was fitted.

Each linear mixed-effects model included a random intercept for subject ID to account for inter-individual heterogeneity of the study population. Fixed effects comprised baseline values of the respective measures (ITA° or melanin index (MIndex) before irradiation), product, ethnicity, product-by-ethnicity interaction, dose, age, sex, and Fitzpatrick skin type. Dose was modelled as a continuous covariate. The effect of dose was assumed to be homogeneous across products and ethnic groups, i.e. no dose-by-product or dose-by-ethnicity interactions were modelled. Sensitivity analyses using alternative model specifications showed that the main results were robust (data not shown).

Models were fitted by restricted maximum likelihood (REML)^25^, which provides stable estimates of variation between subjects. Estimated marginal means (EMMs) and contrasts were derived for the relevant fixed effects. EMMs represent model-based group means adjusted for all covariates, allowing adjusted comparisons between products and ethnicities. Contrasts were calculated to quantify these differences directly. Confidence intervals (95%-CIs) were derived from model coefficients. Two-sided tests were performed at significance level α=0.05.

No imputation of missing outcome data was performed before mixed-effects modelling^26^. One Han Chinese participant discontinued after visit 3. In addition, data from one participant at visit 5 and another at visit 7 were missing because they did not attend the scheduled visits.

## RESULTS

### Study population

Baseline characteristics are summarized in Table 1. The baseline skin pigmentation, expressed as ITA°, was 39.92 ± 10.53 in average across all participants. Participants in the Han Chinese subgroup showed slightly higher ITA° values, indicating lighter baseline pigmentation, though this difference was not statistically significant.

**Table 1.** Baseline characteristics of the study population.

| Characteristics | All subjects<br>(N=40) | Han Chinese<br>(N=20) | Caucasian<br>(N=20) | p-value<br>(Han Chinese vs<br>Caucasian) |
| --- | --- | --- | --- | --- |
| **Sex, n (%)** |  |  |  |  |
| Female | 28 (70.00) | 14 (70.00) | 14 (70.00) | - |
| Male | 12 (30.00) | 6 (30.00) | 6 (30.00) | - |
| **Age, years (mean ± SD)** | $26.65 \pm 5.10$ | $26.30 \pm 4.73$ | $27.00 \pm 5.55$ | 0.839 (NS) |
| **Skin phototypes (Fitzpatrick), n** |  |  |  |  |
| II | 13 | 5 | 8 | - |
| III | 22 | 12 | 10 | - |
| IV | 5 | 3 | 2 | - |
| **Baseline ITA° (mean ± SD)** | $39.92 \pm 10.53$ | $41.83 \pm 10.25$ | $38.00 \pm 10.71$ | 0.254 (NS) |

### Pigmentation responses and product efficacy

The linear mixed-effects models were fitted to assess (i) overall product efficacy across irradiation conditions, and (ii) ethnic differences in product efficacy. A comprehensive summary of effect estimates and p-values for both objectives is provided in the Supporting Information.

### Overall product efficacy

Both products demonstrated significant photoprotective effects after UVA1 exposure across all tanning kinetics (IPD, DT, and PPD; ΔITA°: p<0.001). Reported product effects refer to the calculated contrasts “Control – Product A / B”.

After VL exposure, both products provided significant protection for IPD (ΔITA° - IPD: Product A: -3.48 [95% CI -4.48 to -2.47], p<0.001; Product B: -3.55 [95% CI -4.55 to -2.54], p<0.001), whereas no significant protection was observed for PPD.

Following combined VL *plus* UVA1 exposure, significant protection was again detected for IPD (ΔITA° - IPD: Product A: -2.92 [95% CI -3.90 to -1.93], p<0.001; Product B: -2.27 [95% CI -3.27 to -1.28], p<0.001), while effects on PPD were moderate and statistically non-significant (see Table S1, Supporting Information).

Analyses of changes in MIndex supported these results, showing significant reductions for both products after UVA1 across all tanning kinetics (IPD, PPD, and DT; MIndex: p<0.001). Significant effects were also observed after VL exposure for IPD (VL - IPD: Product A: 0.61 [95% CI 0.19 to 1.03], p=0.005; Product B: 0.65 [95% CI 0.23 to 1.07], p=0.003), and for product B additionally for PPD (VL- PPD: Product B: 0.35 [95% CI 0.07 – 0.63], p=0.014). After combined VL *plus* UVA1 exposure, product A showed a significant effect for IPD (VL+UVA1 - IPD: Product A: 0.48 [95% CI [0.14 to 0.82], p=0.006) (see Table S2, Supporting Information).

### Ethnic differences in product efficacy

In further analyses, interaction effects between ethnicity and product on ΔITA° and ΔMIndex were examined to determine whether sunscreen efficacy differed between Han Chinese and Caucasian participants. Reported product effects refer to the calculated contrasts “Caucasian – Han Chinese”.

A significant interaction between ethnicity and product was detected for ΔITA° after UVA1 exposure, indicating higher efficacy of product A in Han Chinese participants compared to Caucasians for both IPD and PPD (UVA1 – IPD: Product A: -2.45 [95% CI -4.79 to -0.12], p=0.040; UVA1 – PPD: Product A: -2.38 [95% CI -4.60 to -0.17], p=0.035) (see Figure 4 and Table S3 in Supporting Information).

**Figure 4.**
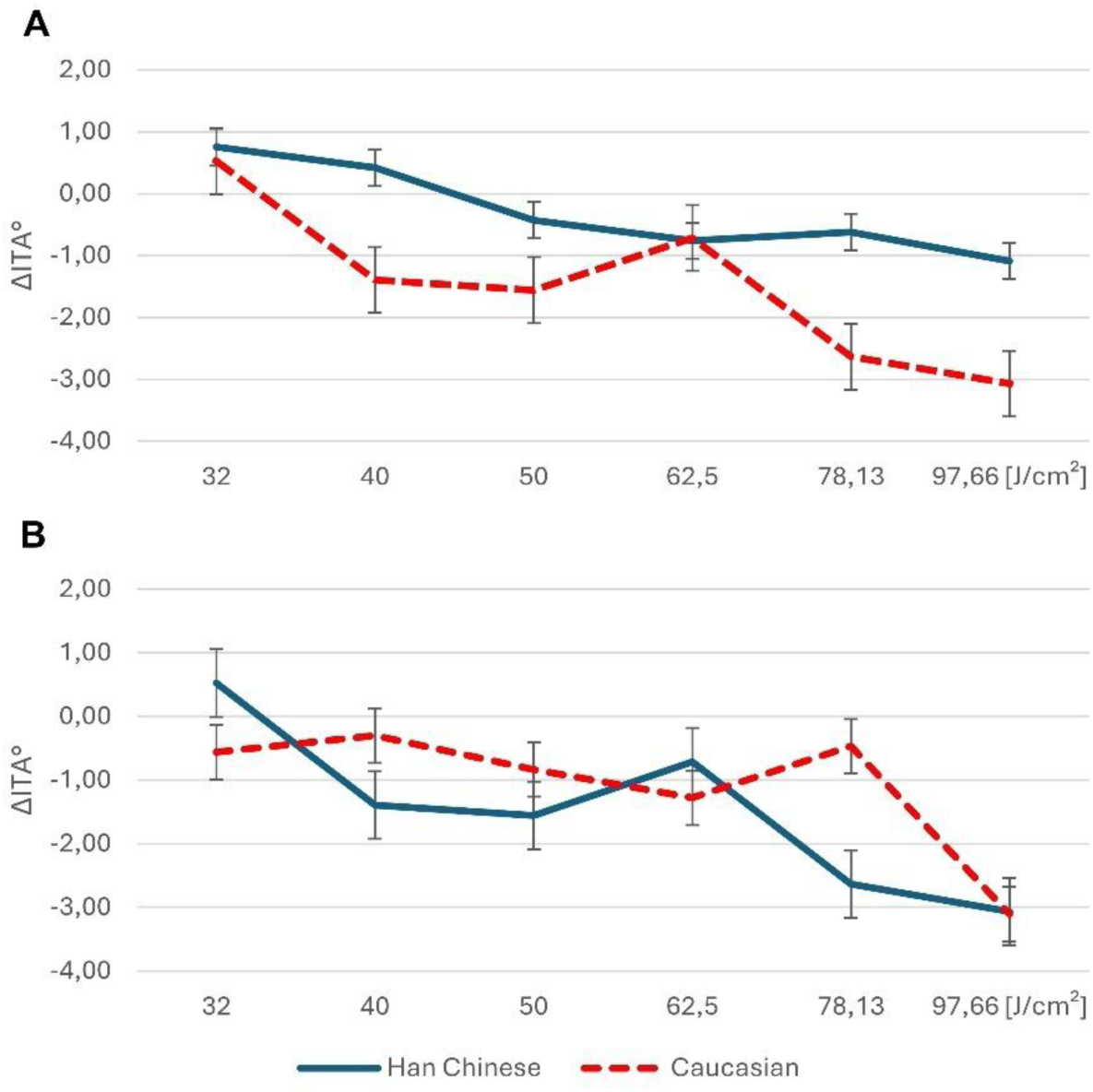
Ethnic differences in dose-dependent changes in persistent pigment darkening (PPD; ΔITA°) after UVA1 exposure for (A) product A and (B) product B. Graphs showing mean changes in Individual Typology Angle (ΔITA°) after exposure to six incremental UVA1 doses in Han Chinese and Caucasian participants. Data are displayed separately for (A) Product A and (B) Product B. Data are presented as means ± standard error of means (SEM).

**Figure 5.**
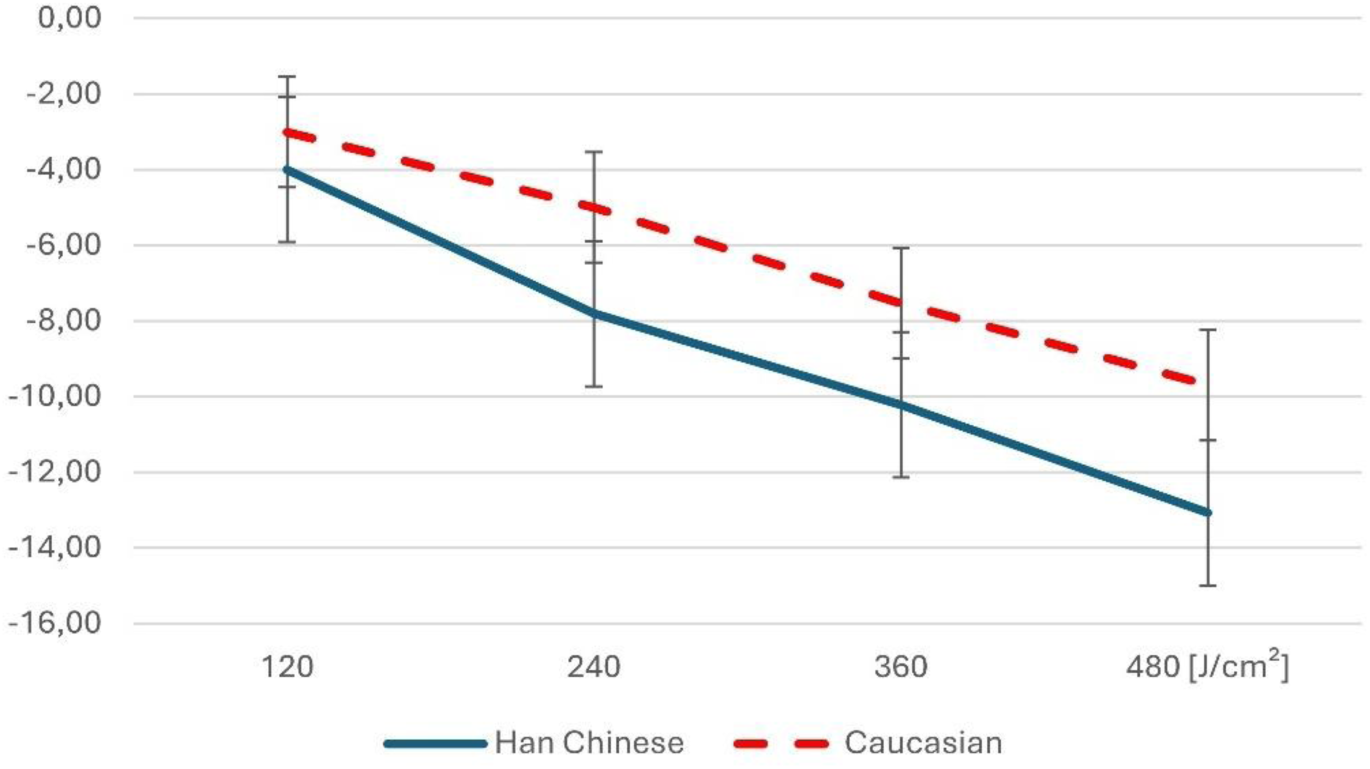
Ethnic differences in dose-dependent changes in persistent pigment darkening (PPD; ΔITA°) in (untreated) control fields after VL *plus* UVA1 exposure. Graphs showing mean changes in Individual Typology Angle (ΔITA°) after exposure to VL plus UVA1 in Han Chinese and Caucasian participants. Data are presented as means ± standard error of means (SEM).

Corresponding ΔMIndex showed consistent but not statistically significant trends (see Table S4 in Supporting Information). In addition, a significant ethnicity-product interaction was observed for MIndex after UVA1 exposure for DT (UVA1-DT: Product A: 1.44 [95% CI 0.17 to 2.71], p=0.027], again indicating higher efficacy of product A in Han Chinese compared to Caucasian participants.

Contrary patterns emerged for VL and combined VL *plus* UVA1 exposures. A significant ethnicity-product interaction for ΔITA° was observed for product B for PPD under both exposure conditions, and for product A for PPD after VL *plus* UVA1 exposure. These results indicated lower product efficacy in Han Chinese (see Table S3 in Supporting Information). Notably, MIndex analysis showed significant ethnicity-product interactions for both product A and B at IPD after combined VL *plus* UVA1 exposure (VL *plus* UVA1 – IPD: Product A: -1.49 [95% CI -2.82 to -0.16], p=0.029; VL *plus* UVA1 – IPD: Product B: -1.54 [95% CI -2.87 to - 0.22], p=0.024), again suggesting reduced efficacy in Han Chinese.

### Ethnic differences in skin pigmentation at unprotected control sites

Across all analyses, a consistent trend towards stronger pigmentation responses in Han Chinese participants was observed after exposure to VL and the combined VL *plus* UVA1 source. None of these differences reached statistical significance. In contrast, after exposure to UVA1 alone, ethnic differences were smaller, non-significant, and showed no consistent pattern. Overall, these findings suggest potential tendency toward greater VL-driven pigmentation in Han Chinese compared to Caucasians (see Table S5 in Supporting Information).

## DISCUSSION AND CONCLUSION

This study evaluated the photoprotective effects of two sunscreen formulations under controlled *in vivo* conditions and compared responses between Han Chinese and Caucasian individuals who were matched for phototype. Both products demonstrated significant protection against pigmentation induced by UVA1, visible light (VL), and combined VL *plus* UVA1, confirming robust efficacy across tested spectral ranges.

Ethnic differences emerged for UVA1-induced immediate (IPD) and persistent pigment darkening (PPD), with product A showing stronger protection in Han Chinese compared to Caucasian participants. Given that baseline pigmentation was comparable between the ethnic groups, these differences are unlikely to be explained by constitutive pigmentation alone.

Although additional ethnicity-related differences in PPD responses were observed after VL *plus* UVA1 exposure, these differences must be interpreted with caution, as neither product showed significant protection against PPD under this combined irradiation. Thus, the observed ethnic differences likely reflect inherent variability in VL-driven pigmentation rather than meaningful differences in sunscreen performance. This interpretation is supported by the observation that Han Chinese participants showed a consistent, although non-significant, trend toward stronger pigmentation after VL and VL *plus* UVA1 exposure across analyses, whereas ethnic differences after UVA1 alone were small and inconsistent.

Both products effectively reduced early pigmentation responses, whereas protection against delayed pigmentation after VL and VL *plus* UVA1 was limited. This aligns with the well-recognized challenge of blocking longer wavelengths, including both initial oxidative processes^27^ and downstream melanogenic signalling^28^, and is consistent with evidence that effective VL protection generally requires tinted sunscreens containing iron oxides^29–32^. Taken together, the results indicate that biological factors beyond phototype contribute to ethnic variation in long-wavelength-induced pigmentation.

A plausible explanation for the stronger UVA1 protection of product A in Han Chinese participants lies in the distinct and superior coverage of long wavelengths achieved through zinc oxide scattering and highly stable UVA1 filters, supported by an oil in water formulation that promotes a homogenous protective film. Product B, although containing a broad set of chemical filters, lacks mineral scattering components and therefore provides less extended attenuation (see Tables S6 and S7 in Supporting Information). The formulations also differ in their antioxidant composition. Under these conditions, the presumed stronger inherent skin responses of Han Chinese skin in the VL range may amplify the observable performance differences.

These findings have practical implications: photoprotection needs may differ even among individuals of similar phototype; ethnicity should be considered when recommending or developing sunscreens, particularly in environments with high UVA1/VL exposure. One possible explanation for these ethnic differences is variation in melanin biology, including differences in eumelanin-to-phaeomelanin ratios, oxidative responsiveness, and wavelength-specific absorption properties, which may alter susceptibility to VL-induced pigmentation. However, these mechanistic assumptions require dedicated investigations. Future studies should focus on developing and validating formulations that target longer wavelengths more effectively. These photoprotection strategies may include the incorporation of mineral filter, optimization of mineral particle size to enhance scattering in the VL-range, and the use of natural derived UV/ VL absorber or antioxidant systems that further mitigate VL-induced oxidative pathways^33,34^.

Limitations of this study include the restriction to young healthy adults, short-term follow-up, and the absence of mechanistic analyses, which limits generalizability and prevents deeper biological interpretation. The modest sample size also reduces the ability to detect small ethnic effects. Future research should define underlying biological drivers and guide the development of sunscreen formulations tailored to diverse skin types and wavelengths-specific photobiological profiles. This could be achieved through controlled in vivo studies combined with mechanistic endpoints (e.g. melanin subtype quantification (eumelanin/ phaeomelanin ratios), or transcriptomic signatures of melanogenesis), as well as comparative studies across phototype-matched ethnic groups to differentiate intrinsic biological variability from product-related effects. In addition, formulation research using standardized VL and VL *plus* UVA1 protection assays – preferably incorporating pigmentary filters such as iron oxides that are known to be effective – may help to identify the technological requirements in these spectral ranges. Together, these approaches would improve our understanding of ethnic differences in VL-induced pigmentation pathways and inform the design of next-generation sunscreens optimized for diverse populations.

In summary, this study provides in vivo evidence that ethnicity – independent of phototype – modulates sunscreen efficacy under UVA1 exposure. These results underscore the concept to consider ethnic background alongside phototype in sunscreen development and clinical recommendations. Furthermore, the observed ethnic variability in skin responses to VL exposures reinforces the need to evaluate VL protection in demographically diverse populations and suggests that VL-targeted formulations may require optimization to ensure effective coverage across diverse populations.

## DATA AVAILABILITY STATEMENT

The data that support the findings of this study are available from the corresponding author upon reasonable request.

