## Supporting Information for "SUNSCREEN EFFICACY AGAINST UVA1- AND VISIBLE LIGHT-INDUCED SKIN PIGMENTATION IS INFLUENCED BY ETHNICITY"

**Table S1. Overall sunscreen efficacy across irradiation conditions (Outcome: ΔITA°)**

| Irradiation condition | Tanning kinetics | Product | Estimate | 95%-CI | p-value |
|---|---|---|---|---|---|
| UVA1 | IPD | A | -6.55 | [-7.14 – -5.96] | **<0.001** |
| | | B | -5.56 | [-6.16 – -4.97] | **<0.001** |
| | PPD | A | -4.81 | [-5.34 – -4.29] | **<0.001** |
| | | B | -4.68 | [-5.20 – -4.15] | **<0.001** |
| | DT | A | -3.47 | [-4.03 – -2.91] | **<0.001** |
| | | B | -3.29 | [-3.85 – -2.73] | **<0.001** |
| VL | IPD | A | -3.48 | [-4.48 – -2.47] | **<0.001** |
| | | B | -3.55 | [-4.55 – -2.54] | **<0.001** |
| | PPD | A | -0.37 | [-1.00 – 0.26] | 0.252 |
| | | B | -0.29 | [-0.92 – 0.34] | 0.370 |
| VL *plus* UVA1 | IPD | A | -2.92 | [-3.90 – -1.93] | **<0.001** |
| | | B | -2.27 | [-3.27 – -1.28] | **<0.001** |
| | PPD | A | -0.61 | [-1.30 – 0.09] | 0.089 |
| | | B | -0.05 | [-0.75 – 0.65] | 0.893 |

*Legend: Assessment of overall product efficacy across all irradiation conditions and the whole study population. Estimates represent contrasts (Control – Product A / B) of estimated marginal means of changes in Individual Typology Angle (ΔITA°) after exposure to different irradiation conditions, analysing different tanning kinetics (IPD, Immediate pigment Darkening; PPD, Persistent Pigment Darkening; DT, Delayed Tanning). Confidence intervals (95%-CI) are presented and statistically significant results (p<0.05) are shown in bold.*

**Table S2. Overall sunscreen efficacy across irradiation conditions (Outcome: ΔMIndex)**

| **Irradiation condition** | **Tanning kinetics** | **Product** | **Estimate** | **95%-CI** | **p-value** |
|---|---|---|---|---|---|
| UVA1 | IPD | A | 3.38 | [3.09 – 3.67] | **<0.001** |
| | | B | 2.72 | [2.42 – 3.01] | **<0.001** |
| | PPD | A | 2.22 | [1.98 – 2.47] | **<0.001** |
| | | B | 2.05 | [1.80 – 2.30] | **<0.001** |
| | DT | A | 1.63 | [1.37 – 1.87] | **<0.001** |
| | | B | 1.37 | [1.12 – 1.62] | **<0.001** |
| VL | IPD | A | 0.61 | [0.19 – 1.03] | **0.005** |
| | | B | 0.65 | [0.23 – 1.07] | **0.003** |
| | PPD | A | 0.16 | [-0.13 – 0.44] | 0.278 |
| | | B | 0.35 | [0.07 – 0.63] | **0.014** |
| VL *plus* UVA1 | IPD | A | 0.48 | [0.14 – 0.82] | **0.006** |
| | | B | 0.34 | [-0.00 – 0.68] | 0.052 |
| | PPD | A | 0.20 | [-0.14 – 0.54] | 0.246 |
| | | B | 0.14 | [-0.20 – 0.48] | 0.415 |

*Legend: Assessment of overall product efficacy across all irradiation conditions and the whole study population. Estimates represent contrasts (Control – Product A / B) of estimated marginal means of changes in melanin index (ΔMIndex) after exposure to different irradiation conditions, analysing different tanning kinetics (IPD, Immediate pigment Darkening; PPD, Persistent Pigment Darkening; DT, Delayed Tanning). Confidence intervals (95%-CI) are presented and statistically significant results (p<0.05) are shown in bold.*

**Table S3. Effects of ethnicity and product on pigmentation (Outcome: ΔITA°)**

| Irradiation condition | Tanning kinetics | Product | Estimate | 95%-CI | p-value |
|---|---|---|---|---|---|
| UVA1 | IPD | A | -2.45 | [-4.79 – -0.12] | **0.040** |
| | | B | -1.44 | [-3.78 – 0.90] | 0.221 |
| | PPD | A | -2.38 | [-4.60 – -0.17] | **0.035** |
| | | B | -1.68 | [-3.89 – 0.53] | 0.133 |
| | DT | A | -2.03 | [-4.18 – 0.12] | 0.063 |
| | | B | -0.30 | [-1.84 – 2.45] | 0.776 |
| VL | IPD | A | 1.59 | [-2.15 – 5.33] | 0.396 |
| | | B | 3.74 | [-0.02 – 7.50] | 0.051 |
| | PPD | A | 1.92 | [-0.34 – 4.18] | 0.093 |
| | | B | 2.34 | [0.07 – 4.61] | **0.044** |
| VL *plus* UVA1 | IPD | A | 1.78 | [-0.87 – 4.44] | 0.184 |
| | | B | 2.05 | [-0.60 – 4.70] | 0.127 |
| | PPD | A | 2.73 | [0.64 – 4.82] | **0.012** |
| | | B | 3.08 | [1.00 – 5.17] | **0.005** |

*Legend: Testing for differences in sunscreen efficacy between Han Chinese and Caucasian participants. Estimates represent contrasts (Caucasian – Han Chinese) of estimated marginal means of changes in Individual Typology Angle (ΔITA°) after exposure to different irradiation conditions, analysing different tanning kinetics (IPD, Immediate pigment Darkening; PPD, Persistent Pigment Darkening; DT, Delayed Tanning). Product A and B refer to the two tested sunscreens. Significant results (p<0.05) are shown in bold. Confidence intervals (95%-CI) are presented.*

**Table S4. Effects of ethnicity and product on pigmentation (Outcome: ΔMIndex)**

| Irradiation condition | Tanning kinetics | Product | Estimate | 95%-CI | p-value |
|---|---|---|---|---|---|
| UVA1 | IPD | A | 1.05 | [-0.17 – 2.27] | 0.089 |
| | | B | 0.98 | [-0.23 – 2.21] | 0.110 |
| | PPD | A | 0.94 | [-0.14 – 2.01] | 0.085 |
| | | B | 0.90 | [-0.18 – 1.97] | 0.100 |
| | DT | A | 1.44 | [0.17 – 2.71] | **0.027** |
| | | B | 0.75 | [-0.52 – 2.01] | 0.242 |
| VL | IPD | A | -0.52 | [-2.15 – 1.10] | 0.519 |
| | | B | -1.21 | [-2.84 – 0.42] | 0.142 |
| | PPD | A | -0.53 | [-1.62 – 0.56] | 0.330 |
| | | B | -0.58 | [-1.67 – 0.52] | 0.293 |
| VL *plus* UVA1 | IPD | A | -1.49 | [-2.82 – -0.16] | **0.029** |
| | | B | -1.54 | [-2.87 – -0.22] | **0.024** |
| | PPD | A | -1.20 | [-2.44 – 0.05] | 0.059 |
| | | B | -1.21 | [-2.45 – 0.03] | 0.056 |

*Legend: Testing for differences in sunscreen efficacy between Han Chinese and Caucasian participants. Estimates represent contrasts (Caucasian – Han Chinese) of estimated marginal means of changes in melanin index (ΔMIndex) after exposure to different irradiation conditions, analysing different tanning kinetics (IPD, Immediate pigment Darkening; PPD, Persistent Pigment Darkening; DT, Delayed Tanning). Product A and B refer to the two tested sunscreens. Significant results (p<0.05) are shown in bold. Confidence intervals (95%-CI) are presented.*

**Table S5. Ethnic differences in pigmentation responses at control sites (Outcome: ΔITA°)**

| Irradiation condition | Tanning kinetics | Estimate | 95%-CI | p-value |
|---|---|---|---|---|
| UVA1 | IPD | -0.60 | [-2.94 – 1.73] | 0.605 |
| | PPD | 0.06 | [-2.15 – 2.27] | 0.954 |
| | DT | 0.86 | [-1.28 – 3.00] | 0.422 |
| VL | IPD | 3.28 | [-0.48 – 7.03] | 0.085 |
| | PPD | 1.63 | [-0.64 – 3.90] | 0.154 |
| VL *plus* UVA1 | IPD | 1.62 | [-1.06 – 4.30] | 0.230 |
| | PPD | 2.01 | [-0.09 – 4.12] | 0.061 |

*Legend: Testing for differences in skin pigmentation between Han Chinese and Caucasian participants using control site data (no product application). Estimates represent contrasts (Caucasian – Han Chinese) of estimated marginal means of changes in Individual Typology Angle (ΔITA°) after exposure to different irradiation conditions, analysing different tanning kinetics (IPD, Immediate pigment Darkening; PPD, Persistent Pigment Darkening; DT, Delayed Tanning). Confidence intervals (95%-CI) are presented.*

**Table S6. Ingredient list (INCI) of product A**

| International Nomenclature of Cosmetic Ingredients (INCI) | |
|---|---|
| AQUA (WATER) | HYDROXYACETOPHENONE |
| CYCLOPENTASILOXANE | STEARIC ACID |
| DISILOXANE | ALUMINUM HYDROXIDE |
| DIMETHICONE | PROPYLENE GLYCOL |
| ZINC OXIDE | ALLANTOIN |
| CAPRYLYL METHICONE | BISABOLOL |
| ETHYLHEXYL SALICYLATE | 1,2-HEXANEDIOL |
| POLYMETHYLSILSESQUIOXANE | CAPRYLYL GLYCOL |
| GLYCERIN | DISTEARDIMONIUM HECTORITE |
| HOMOSALATE | DISODIUM EDTA |
| PHENYLBENZIMIDAZOLE SULFONIC ACID | ORYZA SATIVA (RICE) BRAN EXTRACT |
| DIETHYLAMINO HYDROXYBENZOYL HEXYL BENZOATE | ALUMINA |
| DIETHYLHEXYL BUTAMIDO TRIAZONE | PARFUM (FRAGRANCE) |
| SILICA | ISOPENTYLDIOL |
| TITANIUM DIOXIDE | ECTOIN |
| C12-15 ALKYL BENZOATE | SCUTELLARIA BAICALENSIS ROOT EXTRACT |
| TRIMETHYLSILOXYSILICATE | MALTODEXTRIN |
| TROMETHAMINE | CALOPHYLLUM INOPHYLLUM SEED OIL |
| BIS-ETHYLHEXYLOXYPHENOL METHOXYPHENYL TRIAZINE | CETYL PALMITATE |
| DIBUTYL ADIPATE | CISTUS MONSPELIENSIS EXTRACT |
| MICA | CETEARYL OLIVATE |
| DIMETHICONE/VINYL DIMETHICONE CROSSPOLYMER | ZINGIBER OFFICINALE (GINGER) ROOT EXTRACT |
| DIISOPROPYL SEBACATE | SORBITAN OLIVATE |
| CETYL PEG/PPG-10/1 DIMETHICONE | ETHYLHEXYLGLYCERIN |
| DIMETHYL CAPRAMIDE | NEPHELIUM LAPPACEUM PEEL EXTRACT |
| PEG-10 DIMETHICONE | ASCORBIC ACID POLYPEPTIDE |
| DIPROPYLENE GLYCOL | TOCOPHERYL ACETATE |
| TRIETHOXYCAPRYLYLSILANE | TOCOPHEROL |
| LAURYL PEG-10 TRIS(TRIMETHYLSILOXY)SILYLETHYL DIMETHICONE | CITRIC ACID |

**Table S7. Ingredient list (INCI) of product B**

| International Nomenclature of Cosmetic Ingredients (INCI) | |
|---|---|
| AQUA (WATER) | DECYL GLUCOSIDE |
| OCTOCRYLENE | CARBOMER |
| CYCLOPENTASILOXANE | DISODIUM EDTA |
| HOMOSALATE | PROPYLENE GLYCOL |
| BUTYL METHOXYDIBENZOYLMETHANE | PEG-240/HDI COPOLYMER BIS-DECYLTETRADECETH-20 ETHER |
| ETHYLHEXYL SALICYLATE | TOCOPHERYL ACETATE |
| ISOHEXADECANE | GLYCERIN |
| PHENYLBENZIMIDAZOLE SULFONIC ACID | ORYZA SATIVA (RICE) BRAN EXTRACT |
| SILICA | XANTHAN GUM |
| BUTYLENE GLYCOL | ECTOIN |
| METHYLENE BIS-BENZOTRIAZOLYL TETRAMETHYLBUTYLPHENOL* | SCUTELLARIA BAICALENSIS ROOT EXTRACT |
| DIMETHICONE | MALTODEXTRIN |
| TROMETHAMINE | CALOPHYLLUM INOPHYLLUM SEED OIL |
| DIISOPROPYL SEBACATE | CETYL PALMITATE |
| BIS-ETHYLHEXYLOXYPHENOL METHOXYPHENYL TRIAZINE | CISTUS MONSPELIENSIS EXTRACT |
| POTASSIUM CETYL PHOSPHATE | CETEARYL OLIVATE |
| ETHYLHEXYLGLYCERIN | CAPRYLYL GLYCOL |
| TITANIUM DIOXIDE | SORBITAN OLIVATE |
| HYDROXYACETOPHENONE | ASCORBIC ACID POLYPEPTIDE |
| CETEARYL ALCOHOL | NEPHELIUM LAPPACEUM PEEL EXTRACT |
| DIMETHICONE CROSSPOLYMER | 1,2-HEXANEDIOL |
| ACRYLATES/C10-30 ALKYL ACRYLATE CROSSPOLYMER | POTASSIUM LAURATE |
| GLYCERYL STEARATE | TOCOPHEROL |
| PEG-100 STEARATE | CITRIC ACID |
| SODIUM HYDROXIDE | |

**Figure S1. Irradiation spectra of the light sources used in this study**

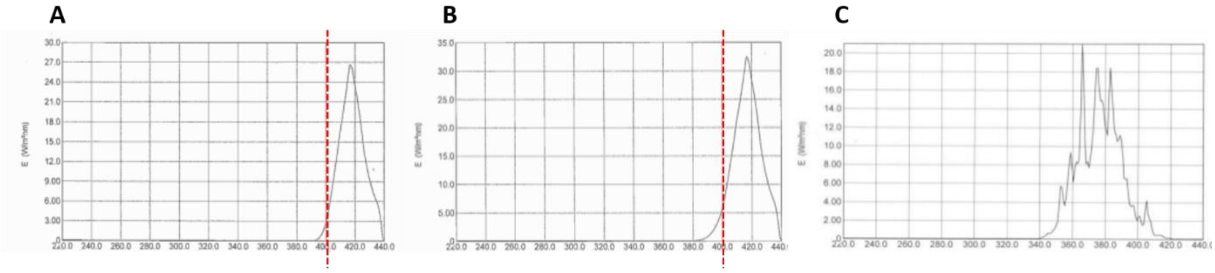


*Legend: (A) Sellas vis400 (visible light: 400 – 440 nm), (B) modified Sellas vis400 (visible light plus UVA1: 390 – 440 nm), and (C) Sellamed 2000 (UVA1: 340 – 400 nm). The red vertical line in panels A and B indicates the spectral differences between VL and VL plus UVA1, marking the additional UVA1 component in the modified vis400 source.*

**Figure S2. Overall photoprotective efficacy of product A and B across all irradiation sources**

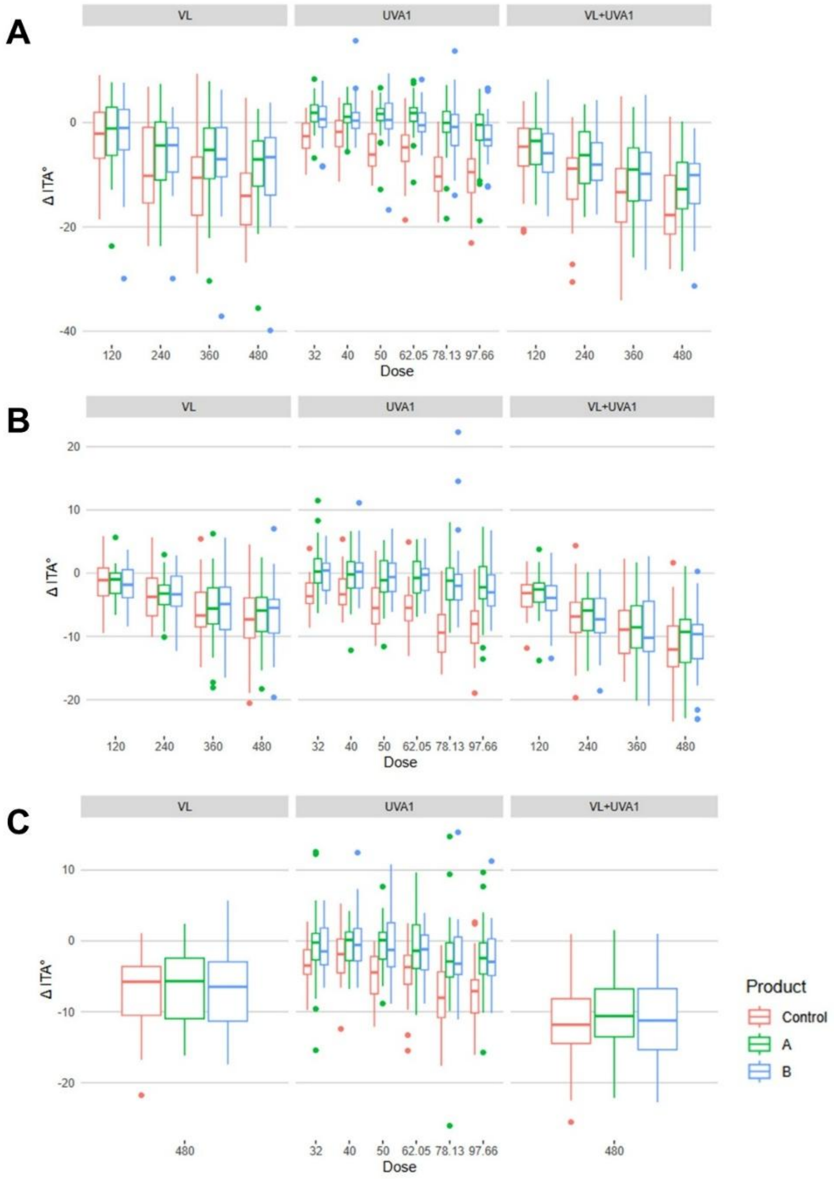


*Legend: Boxplots showing ΔITA° for (A) Immediate Pigment Darkening (IPD), (B) Persistent Pigment Darkening (PPD), and (C) Delayed Tanning (DT) across all tested doses. Each panel compares product A, product B, and control for each irradiation source (visible light (VL), UVA1 and combined VL plus UVA1). For every dose level, the respective ΔITA° values are plotted. Lower ΔITA° indicates stronger pigmentation.*

**Figure S3. Ethnicity-related skin responses to different irradiation sources.**

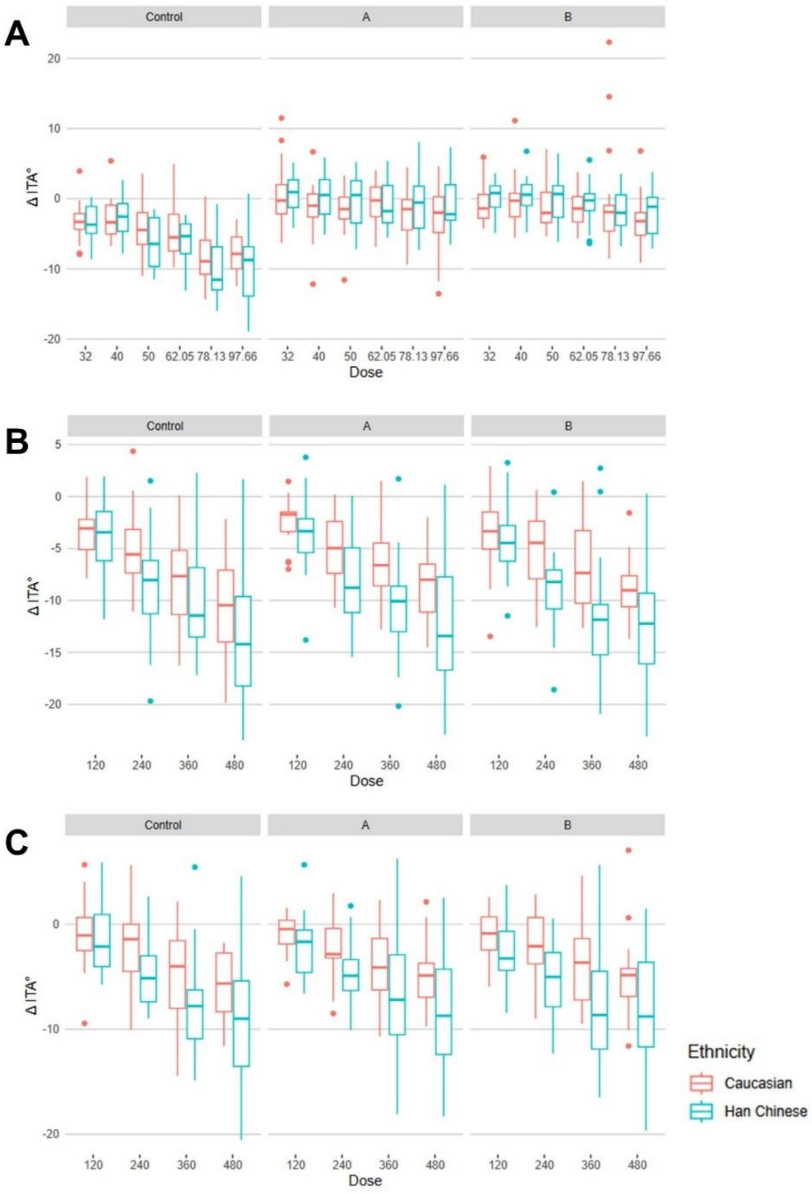


*Legend: Boxplots showing ΔITA° for (A) UVA1, (B) VL plus UVA1, and (C) visible light (VL), comparing skin responses between Han Chinese and Caucasian participants. Each panel displays ΔITA° under control, product A, and product B. conditions. Lower ΔITA° indicates stronger pigmentation.*
